# Effect of Immunoadsorption on clinical presentation and immune alterations in COVID-19–associated ME/CFS

**DOI:** 10.1101/2024.09.25.24314345

**Authors:** Moritz Anft, Lea Wiemers, Kamil Rosiewicz, Adrian Doevelaar, Sarah Skrzypczyk, Julia Kurek, Sviatlana Kaliszczyk, Maximilian Seidel, Ulrik Stervbo, Felix S. Seibert, Timm H. Westhoff, Nina Babel

**Author notes:** Corresponding author: Prof. Nina Babel, Center for Translational Medicine, Medical Department I, Marien Hospital Herne, University Hospital of the Ruhr-University Bochum, Hölkeskampring 40, 44625 Herne, Germany and Berlin Institute of Health, Charité – Universitätsmedizin Berlin, Augustenburger Platz 1, 13353 Berlin, Germany. These authors contributed equally to the manuscript (Moritz Anft, Lea Wiemers, Kamil Rosiewicz).

## Abstract

Autoreactive antibodies (AAB) are currently being investigated as causative or aggravating factors during post-COVID. In this study we analyze the effect of immunoadsorption therapy on symptom improvement and the relationship with immunological parameters in post-COVID patients exhibiting symptoms of Myalgic Encephalomyelitis/Chronic Fatigue Syndrome (ME/CFS). This observational study includes 12 post-COVID patients exhibiting a predominance of ME/CFS symptoms alongside increased concentrations of autonomic nervous system receptors (ANSR) autoantibodies and neurological impairments. We found that following immunoadsorption therapy, the ANSR autoantibodies were nearly eliminated from the patients’ blood. The removal of IgG antibodies was accompanied by a decrease of pro-inflammatory cytokines including IL4, IL2, IL1β, TNF and IL17A serum levels, and a significant reduction of soluble spike protein. Notably, a strong positive correlation between pro-inflammatory cytokines and ASNR-AABs β1, β2, M3, and M4 was observed in spike protein-positive patients, whereas no such correlation was evident in spike protein-negative patients. 30 days post-immunoadsorption therapy, patients exhibited notable improvement in neuropsychological function and a substantial amelioration of hand grip strength was observed. However, neither self-reported symptoms nor scores on ME/CSF questionnaires showed a significant improvement and a rebound of the removed proteins occurring within a month.

## Introduction

While the SARS-CoV-2 pandemic has decreased in intensity over time due to vaccinations and the emergence of less pathogenic virus variants, some patients manifest an array of different symptoms several months after the initial COVID-19 infection. These sequelae are collectively termed Long- or Post-COVID. The detailed research into Post-COVID reveals it to be a heterogeneous clinical picture with over 200 documented different symptoms arising from various causes.^1^ These symptoms are often nonspecific and can impact multiple organs. Alongside cardiovascular symptoms like palpitations or chest pain,^2–4^ patients often report respiratory issues such as breathlessness.^5^ Importantly, neurological and cognitive symptoms including cognitive decline, memory impairment, and fatigue are prevalent among patients experiencing severe Post-COVID effects, leading to significant limitations and even complete immobilization.^6–8^ These symptoms associate with autoantibodies against G protein-coupled receptors (GPCRs) of autonomic nervous system.^9^

Based on current research findings, four main causes can be broadly distinguished as responsible for the array of symptoms.^10^

1. Persistent SARS-CoV-2 virus reservoirs in tissues: Numerous studies have identified SARS-CoV-2 RNA in various tissues,^11,12^ as lung, lymph nodes, and plasma.^13–15^ Additionally, in post-COVID patients, soluble spike protein has been detected in plasma even a year after initial SARS-CoV-2 infection.^16^ It is hypothesized that active, persistent reservoirs of SARS-CoV-2 virus exist in the tissues of a significant portion of post-COVID patients, contributing to the development of subsequent post-COVID symptoms.^17^
2. Autoantibody (AAB) formation:^18^ Infection with SARS-CoV-2 can trigger the formation of autoantibodies targeting components of the immune system, the cardiovascular system, the thyroid gland, as well as rheumatoid-specific autoantibodies and antibodies against G-protein-coupled receptors (GPCR).^19^ Studies show a correlation in the concentration of GPCR AAB with the severity of neurological symptoms.^20^
3. Reactivation of latent viruses: Particularly, the reactivation of various herpes viruses like EBV and HHV-6 has been observed in post-COVID patients, correlating with mitochondrial fragmentation and decreased energy metabolism.^21^
4. Tissue damage from inflammatory processes: Post-COVID patients exhibit elevated levels of pro-inflammatory cytokines such as IL-1ß, IL-6, TNF, or IP10, which can lead to neurological and kidney damage or diabetes mellitus.^22–24^ Additionally, microvascular blood clots with endothelial dysfunction have been detected in a substantial number of patients, associated with symptoms like fatigue, thrombosis, and microclots.^25,26^

The presence of autoantibodies in the majority of post-COVID patients makes immunoadsorption (IA) an attractive therapeutic strategy. There are a variety of different adsorbents, allowing the non-selective removal of all subclasses of immunoglobulins such as IgG or more selective removal of disease-specific molecules such as lipoproteins and CRP. In the context of post-COVID syndrome, IgG immunoadsorption is particularly relevant since it can specifically eliminate autoantibodies. However, preliminary reports on the efficacy of IA in post-COVID are contradictory, rising the need to establish a broader scientific basis.^27–29^

This study aims to explore the impact of IA on neuropsychological impairments in relation to humoral and cellular immune response in post-COVID patients with Myalgic Encephalomyelitis/Chronic Fatigue Syndrome (ME/CFS). The objective is to explore the impact of IA-therapy on diverse markers associated with post-COVID conditions, including autoantibodies directed against autonomic nervous system receptors, soluble spike protein levels, pro-inflammatory cytokines, and the composition of the cellular immune system, both pre- and thirty days after therapy.

## Results

In this study, 12 post-COVID patients with ME/CSF symptoms and high levels of GCPR AAB against autonomic nervous system receptors (ANSR) were evaluated for clinical symptoms, physical and mental health status and immunological characteristics before and after immunoadsorption (IA) therapy. The patients were two-thirds female and with a median age of 50 and a median BMI of 25.25. Among them, 4 out of 12 had ME/CSF developed after SARS-CoV-2 infection, while the remaining 8 described their ME/CSF originally induced by other factors, later aggravated by a subsequent SARS-CoV-2 infection. Table 1 demonstrates patient characteristics and comorbidities. Inclusion criterium was elevated levels of at least 3 of the 4 ANSR proteins β1-adrenergic receptor (>15 U/mL), β2-adrenergic receptor (>8 U/mL), M3-mACh receptor (>6 U/mL), and M4-mACh receptor (>10.7 U/mL). Over a span of 10 days, patients underwent 5 sessions of IA-therapy, with comprehensive screening conducted prior to the first session, after the last session and 30 days after the last session. Health status and symptoms were assessed through detailed anamnesis, while physical examinations encompassed quantification of handgrip strength. Neurological and mental well-being were quantified via the SF-36 questionnaire and various fatigue scores, while dysfunction of the autonomic nervous system was gauged using the COMPASS-31 score. Blood was immunologically analyzed during both assessments and following the last IA session (Figure 1A).

**Figure 1.**
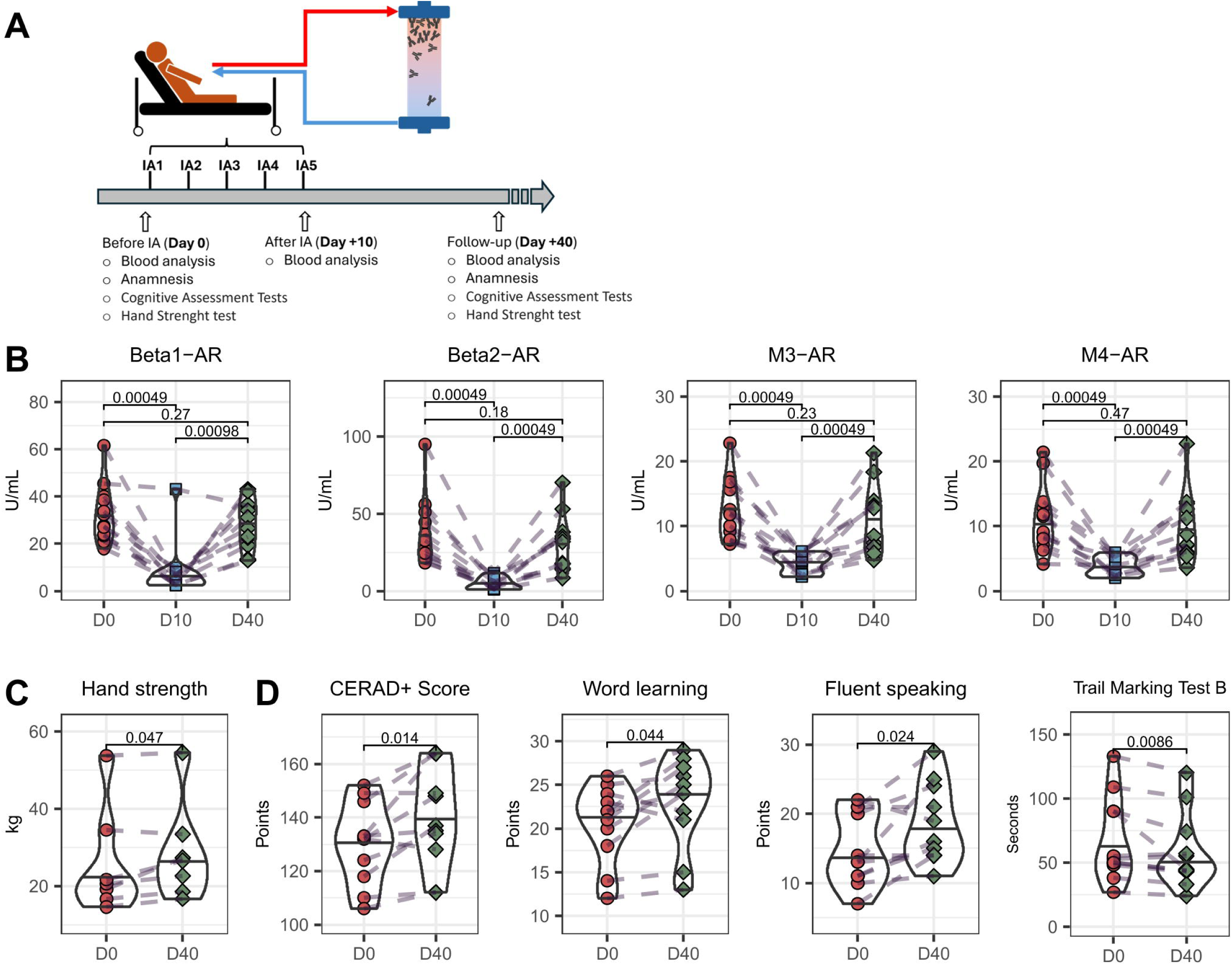
Influence of IA-Therapy on ANSR -AABs and physiological, neuropsychological and cardiological health. (A) Study protocol and time points for various examinations and blood samples. (B) ELISA analysis of the concentration of β1/β2-adrenoceptors and muscarinic M3/M4 receptors before IA (D0), immediately after the last IA session (D10), and 30 days later (D40). (C) Hand strength measurement before and 30 days after the last IA session. (D) Results of the neuropsychological test battery CERAD-Plus before and 30 days after the last IA session. Pairwise Man-Whitney U Test was performed.

**Table 1.** Cohort statistics.

|  |  |
| --- | --- |
| Patients (n) | 12 |
| Age (Median [Q1-Q3]) | 50 [34.5-60] |
| Sex (Female; n (%)) | 8 (66.7%) |
| BMI (Median [Q1-Q3]) | 25.25 [22.1-26.9] |
| ME/CSF | 12 (100%) |
| - Trigger SARS-CoV-2 infection | 4 (33.3%) |
| - Other trigger, worsened by SARS-CoV-2 infection | 8 (66.6%) |
| Comorbidities |  |
| Arterial hypertension (n (%)) | 3 (25.0%) |
| Hyperlipidemia (n (%)) | 3 (25.0%) |
| COPD (n (%)) | 2 (16.7%) |
| Nicotine abuse (n (%)) | 2 (16.7%) |
| ESRD (n (%)) | 0 (0%) |
| Diabetes mellitus (n (%)) | 0 (0%) |

BMI = Body mass index; COPD = chronic obstructive pulmonary disease; ESRD = End stage renal disease

### IA-therapy leads to a strong decrease in ANSR-AABs and to an improvement in multiple physical and neuropsychological factors

Quantifying the ANSR autoantibodies in the patients’ serum immediately after the final therapy session revealed that the levels of autoantibodies was significantly reduced below their respective threshold values through IA. 30 days later, however, the concentration of autoantibodies had returned to a slightly, but not significantly, lower level compared to the time before therapy (Figure 1B). 30 days after IA-therapy, the patients had median autoantibody levels of 93.9% (β1), 74.8% (β2), 82.4% (M3), and 88.8% (M4) compared of the baseline levels before IA (Supplemental Table S1). In addition, several physical and neuropsychological were observed following the IA. Patients exhibited significantly increased hand strength post-therapy (Figure 1C). Moreover, the overall score of the CERAD test battery was significantly higher at the one-month follow-up measurement, primarily attributed to enhancements in word learning, fluent speaking, and the Trail Making Test B (Figure 1D).

### IA-therapy does not lead to self-perceived changes

In addition to these objective measurements, several questionnaires and anamnesis were used to assess patients’ self-perception of whether and how their symptoms and physical and mental health changed after the therapy (Table 2). The surveys revealed no significant differences in the perception of physical and mental health following the IA-therapy. Neither in the SF-36 questionnaire regarding health-related quality of life nor in the Chandler Fatigue Score, Bell Score, or the Canadian criteria for ME/CFS showed a significant improvement or deterioration due to the therapy (Table 2 – upper section). Additionally, the self-reported symptoms of the patients did not differ 30 days after the last IA compared to before the therapy (Table 2 – lower section).

**Table 2.** Reported symptoms, and self-rated physical and mental health.

| Test | Parameter | Before IA | Follow-Up month) | (1 P-Value |
| --- | --- | --- | --- | --- |
| **Self rated physical and mental health** |  |  |  |  |
| **SF-36 questionnaire** |  |  |  |  |
|  | **SF36-Total Score** (Mean (SD)) | 29,7 (7.2) | 31,7 (11.9) | 0.57<sup>¥</sup> |
|  | **Physical health** |  |  |  |
|  | Physical functioning (Mean (SD)) | 29,2 (16.2) | 29,1 (18.8) | 0.67<sup>¥</sup> |
|  | Role-physical (Mean (SD)) | 8,3 (15.6) | 2,3 (7.2) | 0.37<sup>¥</sup> |
|  | **Mental health** |  |  |  |
|  | Mental health (Mean (SD)) | 51,7 (14.1) | 60,7 (16.7) | 0.12<sup>¥</sup> |
|  | Vitality (Mean (SD)) | 13,9 (13.19) | 15,9 (12) | 1.00<sup>¥</sup> |
|  | Role-emotional (Mean (SD)) | 66,6 (43.1) | 81,8 (38.6) | 0.41<sup>¥</sup> |
|  | Social functioning (Mean (SD)) | 13,4 (14) | 17 (15.3) | 0.31<sup>¥</sup> |
|  | General health (Mean (SD)) | 22 ± (2) | 22,7 (12.3) | 0.72<sup>¥</sup> |
|  | Bodily pain (Mean (SD)) | 28,6 (27) | 36,2 (25.5) | 0.09<sup>¥</sup> |
| **Other questionnaires** |  |  |  |  |
|  | Chalder Fatigue Scale (bm) (Mean (SD)) | 10.3 (0.9) | 9.2 (2.2) | 0.28<sup>¥</sup> |
|  | Bell Score (Mean (SD)) | 30 (7.4) | 31.8 (11.1) | 0.57<sup>¥</sup> |
|  | Canadian criteria for ME/CFS (n (%)) | 10/11 (90.9%) | 9/11 (81.8%) | 1.00* |
|  | neurological/cognitive manifestations (n (%)) | 11/11 (100%) | 10/11 (90.9%) | 1.00* |
|  | Immunological manifestations (n (%)) | 11/11 (100%) | 11/11 (100%) | 1.00* |
| **Self reported symptoms** |  |  |  |  |
|  | Flu-like symptoms (n (%)) | 10/11 (90.9%) | 10/11 (90.9%) | 1.00* |
|  | Persistent cough (n (%)) | 1/11 (9.1%) | 0/11 (0%) | 1.00* |
|  | Dyspnoe (n (%)) | 10/11 (90.9%) | 8/11 (72.7%) | 0.57* |
|  | Tiredness (n (%)) | 11/11 (100%) | 11/11 (100%) | 1.00* |
|  | Respiratory pain (n (%)) | 0/11 (0%) | 1/11 (9.1%) | 1.00* |
|  | Thoracic Pain (n (%)) | 2/11 (18.2%) | 1/11 (9.1%) | 1.00* |
|  | Odor changes (n (%)) | 2/11 (18.2%) | 1/11 (9.1%) | 1.00* |
|  | Changes in taste (n (%)) | 2/11 (18.2%) | 1/11 (9.1%) | 1.00* |
|  | Rhinorrhoe (n (%)) | 5/11 (45.5%) | 6/11 (54.5%) | 1.00* |
|  | Cephalgien (n (%)) | 10/11 (90.9%) | 11/11 (100%) | 1.00* |
|  | Myalgien (n (%)) | 8/11 (72.7%) | 7/11 (63.6%) | 1.00* |
|  | Limb pain (n (%)) | 8/11 (72.7%) | 8/11 (72.7%) | 1.00* |
|  | Unusual abdominal pain (n (%)) | 4/11 (36.4%) | 4/11 (36.4%) | 1.00* |
|  | Nausea (n (%)) | 2/11 (18.2%) | 0/11 (0%) | 0.47* |
|  | Diarrhoea (n (%)) | 3/11 (27.3%) | 1/11 (9.1%) | 0.59* |
|  | Confusion, disorientation or drowsiness (n (%)) | 4/11 (36.4%) | 2/11 (18.2%) | 0.64* |
|  | Brain Fog (n (%)) | 10/11 (90.9%) | 11/11 (100%) | 1.00* |
|  | Sleep disorders (n (%)) | 9/11 (81.8%) | 9/11 (81.8%) | 1.00* |
|  | Lack of drive (n (%)) | 3/11 (27.3%) | 2/11 (18.2%) | 1.00* |

SD = standard deviation; bm = bimodular; \* = Fisher's Exact Test, ¥ = Mann Whitney
U Test

### Temporary reduction in pro-inflammatory cytokines

The concentration of pro-inflammatory cytokines in the serum of patients was examined before and after the IA. Especially IL-4, IL-2, IL-1β, TNF and IL-17A were significantly reduced immediately after the IA compared to the pre-therapy level. Similar to the concentration ANSR AABs, the concentration of all these cytokines rose back to a similar level as before the IA-therapy in the follow-up analysis 30 days later. Interestingly, the concentration of TGFβ1 increased significantly directly after IA and dropped back to the pre-IA level in the follow-up analysis, and the concentration of IL-8 was significantly reduced after IA-therapy (Figure 2). The concentration of other cytokines such as IP-10, MCP-1, IL-6 and IL-10 were not changed by the therapy (Supplemental Figure S1). In addition, 5 of the 12 patients had an acute EBV infection with an elevated EBV load during the whole study, and the viral load did not change during and after IA-therapy (Supplementary Figure S2).

**Figure 2.**
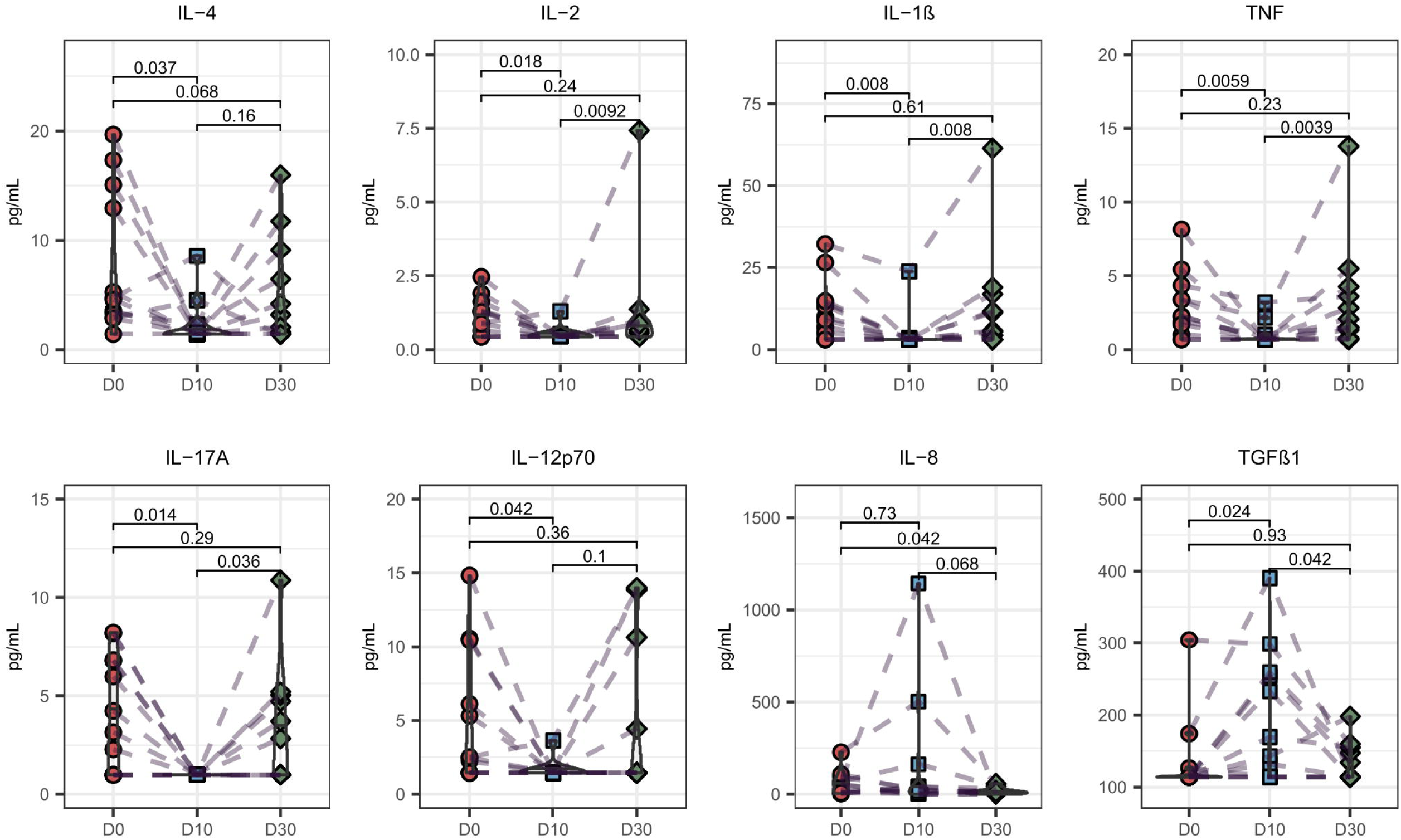
Temporary reduction of pro-inflammatory cytokines after IA-Therapy. Analysis of the cytokine profile of IA patients before IA, immediately after IA and 30 days after the last IA. Pairwise Man-Whitney U Test was performed.

### Cellular immunity is unaffected by IA

To explore if the perturbations in pro-inflammatory cytokines during IA-therapy had any effect on the composition of T cell populations, the major T cell populations before and 30 days after IA-therapy were examined. While there were no differences in the percentage of total CD4^+^ and CD8^+^ T cells, patients demonstrated significantly more naive and significantly fewer memory CD4^+^ T cells and additionally significantly fewer CXCR5 expressing follicular helper cells in the follow-up examination compared to the data before the start of IA-therapy (Figure 3A+3C).

**Figure 3.**
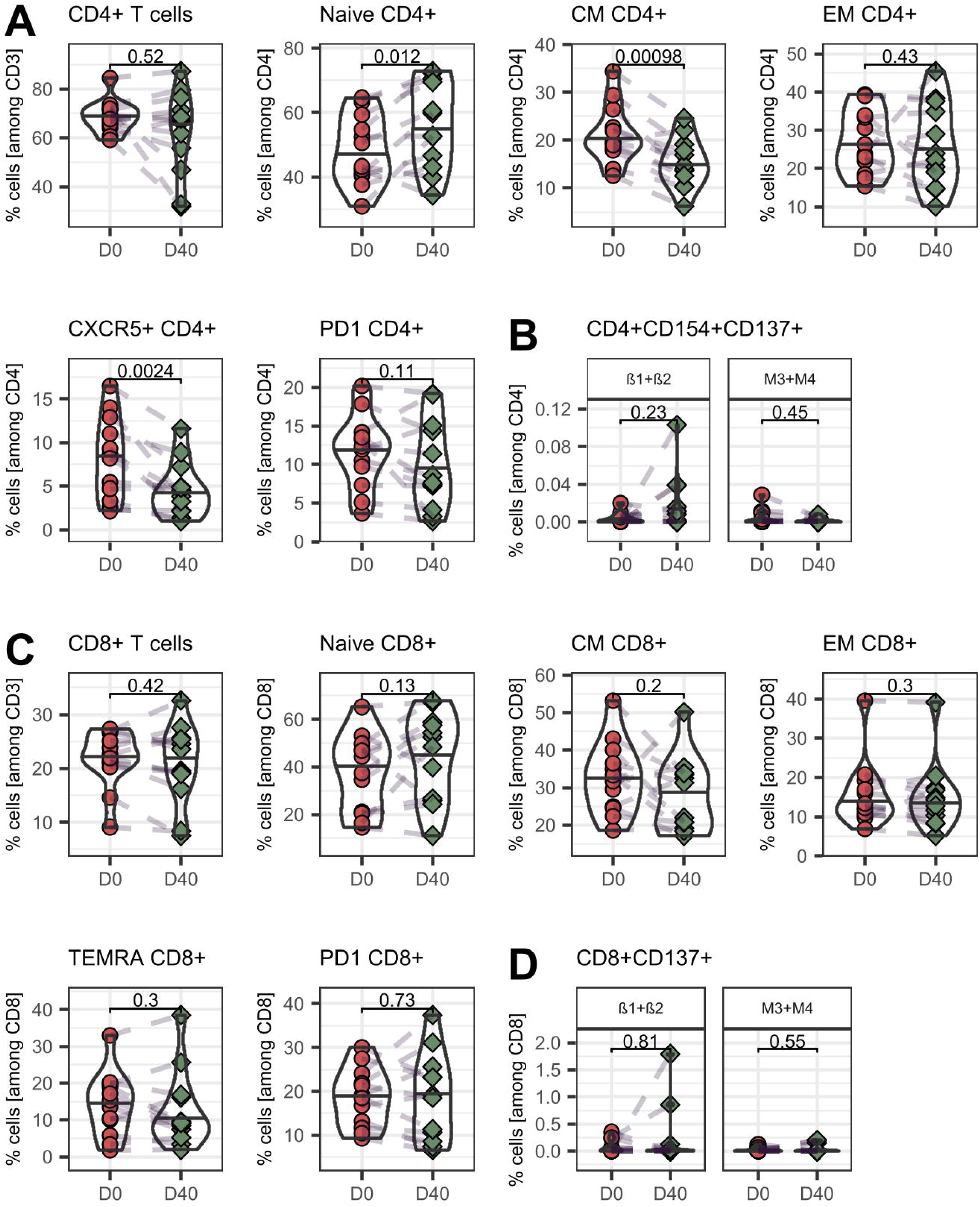
Percentage of T cell populations and ANSR-specific T cells. PBMCs from patients were isolated before IA (D0), and 30 days after the last IA (D40) and stained with the appropriate antibodies and analyzed in a flow-cytometer. (A+C) T cells were identified as CD3^+^ and CD4^+^ or CD8^+^. Naïve and memory T cells were distinguished by expression of CD45RA and CCR7, follicular helper cells by CXCR5 and exhausted T cells by expression of PD-1. CM = Central Memory, EM = Effector Memory. (B+D) PBMC were stimulate for 18h with SARS-CoV-2 Spike OPPs, β1/β2-adrenoreceptors or muscarinic M3/M4 receptor cell lysates, stained with the appropriate antibodies and analyzed in a flow-cytometer. Activated T helper cells were identified as CD4^+^ CD154^+^CD137^+^ and activated cytotoxic T cells as CD8^+^CD137^+^. Pairwise Man-Whitney U Test was performed. Gating strategies are depicted in supplementary figure S5+S6.

T cells may become exhausted in a prolonged pro-inflammatory.^30^ We therefore explored whether this is the case in post-COVID before and after IA-therapy. The frequency of PD-1 expression on CD4^+^ and CD8^+^ T-cells prior to the therapy was within previously observed ranges (Figure 3A+C)^31^. Except for a significant decrease in Tim-3 expressing CD4 T cells, there were no differences in the expression of exhaustion markers PD-1, Tim-3, CTLA-4, or Lag-3 directly or 30 days after IA (Figure 3 A+C and Supplemental Figure S3).

To investigate if the autoimmunity towards autonomic nervous system receptors extended to the T cells, PBMC were stimulated with pooled lysates of β1/β2-adrenoceptors and muscarinic M3/M4 receptors, respectively (Figure 3B+D). Simultaneously, the potential impact on cellular immunity to SARS-CoV-2 was examined by additionally stimulating of PBMC with a pool of SARS-CoV-2 spike protein overlapping peptides (Figure 3B+D). ANSR specific T cells were quantified by assessing the expression of CD137 on CD8^+^ T cells and CD137 and CD154 on CD4^+^ T cells. Following IA-therapy, there were no discernible differences in the percentages of CD4 and CD8 T cells specific to β1/β2-adrenoceptors or muscarinic M3/M4 receptors compared to the respective frequencies before IA-therapy. Moreover, although a slight yet significant decrease was noted specifically in the proportion of naive CD4 T cells targeting M3+M4 after IA, no significant differences were observed in the proportions of naive, memory, and exhausted ANSR-specific T cells post-IA (Figure 3B+D and supplemental Figure S4).

### IA-Therapy leads to a temporary reduction of SARS-CoV-2 spike protein

Several studies have shown that a subset of post-COVID patients have detectable levels of soluble spike protein in their blood.^16,17^ The next step was to investigate the development of spike protein levels in the patients in this study before the IA-therapy, immediately after the last IA and at follow-up. Before the first IA, soluble spike protein was detected in 6 out of 12 patients, with a median concentration of 207.7 pg/mL. Following the IA-therapy, the concentration of soluble spike protein significantly dropped to 22.8 pg/mL, but rebounded to a median of 207.5 pg/mL by the 30-day follow-up examination (Figure 4A). Except for one patient, the IA-therapy also significantly reduced the concentration of SARS-CoV-2 spike protein specific antibodies, but these levels also returned to their initial values within the 30 days following IA (Figure 4B). However, we found no differences in the frequency of spike protein specific CD4^+^ or CD8^+^ T cells after the IA-therapy (Figure 4C).

**Figure 4.**
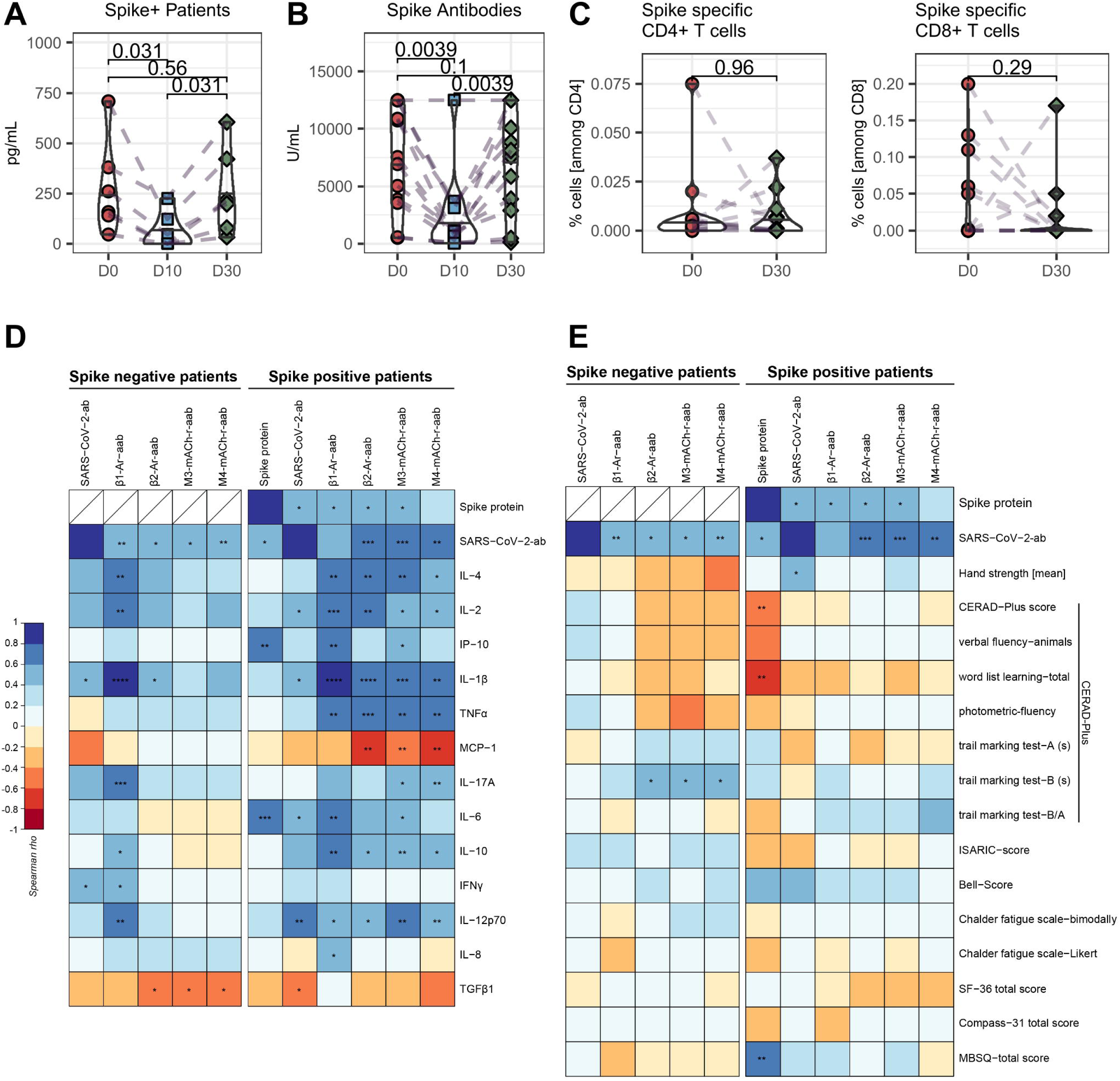
Some of the patients have soluble spike protein, which is correlated with ANSR AAB and blood levels of cytokines. (A) Concentration of spike protein in all spike positive patients (6/12). (B) Concentration of serum antibodies against SARS-CoV-2 spike protein. (C) Percentage of spike protein specific T cells. PBMC were stimulated for 18h with a SARS-CoV-2 spike protein overlapping peptide pool. Spike specific T helper cells were identified as CD4^+^CD154^+^CD137^+^ and spike specific cytotoxic T cells as CD8^+^CD137^+^ among all CD4 or CD8 T cells. (D+E) Correlation matrices of spike+ and spike-patients with ANSR AABs vs. blood cytokines (D) and neurological test (E). All collected values were tested for correlation. *P*-values are shown as asterisks within the boxes. \**p*< 0.05, \*\**p*< 0.01, \*\*\**p<* 0.001, \*\*\*\**p*< 0.0001. Color grading refers to Spearman’s rho rank correlation and is indicated as legend underneath the correlation matrix.

### Correlation between neurological symptoms, virological and immunological status

As stated above, the ratio of patients identified with soluble spike protein at study start was 50:50, whereas 100% of patients showed detectable SARS-CoV-2 specific antibodies. This interesting result led to the question, if there are possible differences between these two groups based on the detected soluble spike protein (groups named as “spike-” and “spike+”). First, spike+ patients were characterized by significantly strong positive correlations between spike+ protein and SARS-CoV-2 specific antibodies, ANSR autoantibodies (β1, β2 and M3) and pro-inflammatory cytokines IP-10 and IL-6 (Figure 4D). SARS-CoV-2 specific antibodies showed not only nearly the same significant correlation with ANSR autoantibodies (β2, M3 and M4) but also with pro-inflammatory cytokines IL-2, IL-1β, IL-6 and IL-12p70 and a significant negative correlation to the anti-inflammatory cytokine TGFβ1. In general, all ANSR autoantibodies showed very strong correlations to all cytokines except for MCP-1, INFγ and TGFβ1. This result was in strong contrast to spike-patients. Here, SARS-CoV-2 spike protein specific antibody levels correlated only positively with blood levels of IL-1β and INFγ. Interestingly, ANSR autoantibody levels (β2, M3, M4) showed a significant negative correlation to TGFβ1 blood levels. Only the positive correlation between ANSR autoantibody levels and SARS-CoV-2 specific antibody levels had spike- and spike+ patients in common (Figure 4D).

Next, the detected levels of soluble spike protein, SARS-CoV-2 specific antibodies and ANSR autoantibodies were correlated with the monitored clinical ME/CSF data. Again, spike+ patients showed a slightly different correlation profile than spike-patients. Soluble spike protein levels correlated significantly positive with MBSQ total score, while significantly negative with CERAD-Plus score and CERAD-Plus (word list learning) (Figure 4E).

This study shows that TheraSorb IA-therapy has the potential to improve the neuropsychological state of ME/CFS, but has no positive impact on self-perceived symptoms and mental health. In addition to ANSR-AABs, it effectively removes the soluble spike protein, decreases pro-inflammatory cytokines and increase inflammatory cytokine TGFβ. However, these improvements are highly transient, with a resurgence of the eliminated proteins happening within a month. Further therapeutic steps are needed to be implemented to provide a long-term immunological and symptomatic improvements

## Discussion

Although the SARS-CoV-2 pandemic is over, humankind is still struggling with its consequences years later. The occurrence of post-COVID after SARS-CoV-2 infections poses major challenges for both patients and healthcare providers. The multiple symptoms and underlying causes make diagnosis and treatment complex and difficult. ANSR autoantibodies are associated with disease severity in ME/CSF and their removal is associated with moderate improvement in symptoms in a previous study.^28^ The present study shows that the health status of post-COVID patients with ME/CFS symptoms and high ANSR autoantibody levels objectively improved after IA-therapy. However, the affected patients did not perceive any improvement as reflected in the self-reported symptoms.

In the present study, all patients had high titers of ANSR autoantibodies against β1/β2-adrenoceptor and muscarinic M3/M4 receptor. These ANSR and other GPCR autoantibodies were earlier associated with the occurrence and worsening of multiple diseases like cardiovascular or rheumatic diseases,^32–34^ and several studies have shown a correlation between the presence of these ANSR autoantibodies and the severity of post-COVID.^9,35–37^ In the context of ME/CSF and post-COVID, it has been shown that ANSR autoantibodies may be associated with Postural Orthostatic Tachycardia Syndrome (POTS).^38,39^ It is suggested that these ANSR autoantibodies can disrupt autonomic nervous system function by interfering with signaling pathways through binding to ANSRs, leading to symptoms such as excessive increase in heart rate upon standing, dizziness, and fatigue.^40^ The patients in this study also reported these neurological and hemodynamic symptoms and were diagnosed with fatigue. Self-assessments using the SF-36 questionnaire revealed substantial deficits in both physical and mental health. Additionally, the Bell and Chalder Fatigue Scores were low, consistent with findings from studies on post-COVID patients with ME/CFS symptoms.^41^ IA-therapy could effectively remove ANSR autoantibodies in patients with ME/CSF and provided a moderate improvement in symptoms in the period shortly after therapy.^28,42^ In line with this, an overall score increase in the CERAD+ neuropsychological test as well as an increase in hand strength after removal of the ANSR autoantibodies in this study was observed.

As also reported in other studies,^28,42^ the levels of autoantibodies returned to their original levels one month after therapy, which could be the reason that in contrast to the improvement of objective symptoms, there was no demonstrated improvement in the quantified fatigue scores and self-reported symptoms by the patients in the follow-up. While neurological symptoms such as tiredness, cephalgia and brain fog were present in almost all patients before therapy, they did not improve by IA-therapy and the removal of ANSR autoantibodies. Accordingly, physical and mental health were at the same low level after IA-therapy as before treatment, suggesting that short-term removal of these AABs from the blood by IA-therapy cannot effectively treat post-COVID symptoms. In contrast, removal of ANSR autoantibodies in a post-COVID patient using the DNA aptamer BC 007 resulted in a significant and four-week lasting improvement in symptoms such as fatigue symptoms, taste and capillary microcirculation^43^. It is possible that a long-lasting DNA aptamer neutralizes ANSR autoantibodies over an extended period, rather than only targeting the antibodies present during the therapy, as is the case with IA-therapy. Moreover, IA-therapy can only filter AABs out of the blood, whereas DNA aptamers can also act at the tissue level.^44^

Interestingly, changes in pro-inflammatory cytokine levels were detected after the IA-therapy. While some cytokines, such as IL-2, IL-4, and IL-17A, were reduced immediately after the final IA-therapy TGFβ1 levels were elevated. Other cytokines, like IP-10 and MCP-1, were not significantly affected by the therapy. The reason for the changes in cytokine levels after IA remains speculative. The different impact of IA on certain cytokines demonstrates that this is not a pure mechanical cytokine elimination. There might be a general reduction in inflammation after IA, as autoantibodies drive inflammation via the activation of the complement system or direct antibody-dependent activation of immune cells.^45–47^ Furthermore, patients with infectious or autoimmune diseases have autoantibodies against some cytokines that bind to the IA column, which could also remove bound cytokines.^48–50^

While the presence of ANSR autoantibodies indicate a defective immunotolerance after a viral infection,^51,52^ the existence of soluble spike protein in post-COVID patients may indicate an active, persistent infection with SARS-CoV-2 viruses as it is currently discussed. Persistent viral reservoirs are known from other RNA viruses such as Ebola or Measles and are found in various tissues.^53,54^ The presence of soluble SARS-CoV-2 spike protein in 6 of the 12 patients suggests that half of the post-COVID patients in this study contain active virus reservoirs in their tissues that release spike protein into the bloodstream. Interestingly, spike protein was also largely removed from the patients’ blood with IA-therapy, even though they were not expected to bind to the Ig-column. However, since high titers of SARS-CoV-2 spike protein specific antibodies were detected in all patients, it is likely that these antibodies bound to soluble spike protein, leading to their removal along with the antibodies during IA-therapy. Notable, there was no improvement in self-reported symptoms after IA-therapy associated with the removal of spike proteins in patients with soluble spike protein, which is consistent with the observation that spike positive patients do not show significant correlations between spike level and ME/CSF test scores. Whether and how soluble spike protein itself has a direct pathogenic effect or if the active virus reservoirs lead to the manifestation and exacerbation of post-COVID symptoms is not yet clear. It is discussed that soluble spike protein directly leads to the formation of fibrin and amolytic microclots, which should be prevented by removing spike protein from the body.^55^ Additionally, it has been shown in mice that spike protein injected in the brain can lead to neuropsychological impairments that are similar to post-COVID symptoms,^56^ although it remains highly unclear, whether the spike protein crosses the blood-brain barrier in humans and can be responsible for similar mechanisms. However, since IA-therapy does not address the source of the soluble spike protein, the ongoing production of spike protein due to persistent infection may prevent a beneficial effect. This would be supported by the fact that the level of spike protein almost returned to its original level shortly after the last IA session. It is also noteworthy that a strong correlation between the amount of all ANSR autoantibodies tested and most pro-inflammatory cytokines was only seen in the spike positive patients, which is only the case for β1 AAB in the spike negative patients. It could be assumed that a persistent infection leads to increased inflammation and generation of auto-reactive antibodies. Moreover, previous studies demonstrated a clear association between inflammation and autoimmunity.^57^

Also some changes at the cellular level were observed after IA-therapy. There was a significant reduction in CD4^+^ central memory and follicular helper T cells, accompanied by an increase in the frequency of naive T cells. A direct interaction and removal of the cells by the Ig-binding apheresis column can be excluded due to the high specificity of the column.^58^ However, the IA procedure might lead to a temporary redistribution of T cell subsets within the body. The stress associated with the treatment could trigger a short-term migration of T cells from the bloodstream to various tissues, causing an apparent reduction of these cells in the peripheral blood. Additionally, the observed changes in the cytokine profile, which is fundamental for T cell homeostasis, could impact the maintenance of these T cell populations and result in alterations in the distribution of sub-populations.^59^ One difficulty with persistent and chronic infections is that ongoing activation induced by constant antigen presence can lead to exhaustion and changes in the differentiation of T cells.^60,61^ Consequently, it was shown that T cells from post-COVID patients express higher levels of exhaustion markers PD-1, CTLA-4 and Tim-3.^62,63^ Although this study did not compare the levels of exhaustion markers or the T cell frequencies between post-COVID patients and convalescent controls, any differences in the frequency of ANSR lysate-specific or spike protein-specific T cells and their PD-1, CTLA-4 and Tim-3 expressing T cell subsets before and one month after IA-therapy have been observed. These data suggest that even though these antigens were almost completely removed immediately after therapy, IA-therapy has a short effect on these molecules and obviously no modulating effect on T cells.

The fact that there was no improvement in the patients’ perception of their symptoms may also be related to the IA procedure itself. IA-therapy is an invasive procedure that poses a challenge for all patients due to the long duration and use of a Shaldon catheters. Especially for patients in a poor general condition, such as post-COVID patients, this can further increase the level of suffering. In these cases, subjectively reported symptoms may not align with objective measurements

This study has some limitations. This is an observational study, and a randomized control trial including a placebo group is needed to verify the beneficial effect of IA-therapy. Furthermore, the small sample size of 12 patients reduces the statistical power, potentially masking significant changes. Fortunately, much of the laboratory data is consistent enough among patients to allow reasonably firm conclusions. However, the study does not prove a causal relation between ANSR autoantibodies and ME/CFS symptoms, since IA removes other immunoglobulins as well. Another limitation is a potential practicing effect in the neuropsychological tests. However, it has been shown that these can be eliminated or limited by the selection of tests^64^. The follow up at day 30 after IA is relatively short, and it will be necessary to see if the observed improvements are persistent and even lead to a self-assessed improvement.

In summary, this study demonstrates that IA-therapy in patients with post-COVID disease and ME/CSF symptoms leads to a significant reduction in autoantibodies, soluble spike protein, and inflammatory cytokines. This is accompanied by moderate improvements in neuropsychological parameters, though it does not influence the patients’ subjective perception of their symptoms. These data lay the foundation for further research to determine the extent of these observed benefits for patients and whether this treatment should be considered standard for post-COVID care in the future.

## Methods

### Study population and design

This observational study involved 12 post-COVID patients exhibiting a predominance of ME/CFS symptoms alongside the presence of ANSR autoantibodies (β1 > 15 U/mL; β2 > 8 U/mL; M3 > 6 U/mL; M4 > 10.7 U/mL) and neurological impairments. Age over 18 years, ME/CSF syndrome, the presence of 3 of the 4 ANSR AABs and written consent were defined as inclusion criteria. Patients underwent immunoadsorption therapy and blood samples were collected from patients before the initial session, immediately post-treatment, and again after an additional 30 days (Figure 1). The study was approved by the ethical committee of Ruhr university Bochum (Reg.-Nr. 23-7752) and all participants provided a written informed consents.

### Immunoadsorption Therapy

Patients underwent a course of TheraSorb - Ig omni 5 adsorber (Miltenyi, Germany) immunoadsorption therapy comprising 5 sessions over a span of 10 days aimed at eliminating IgG antibodies. The therapy was carried out at Marien Hospital Herne in Medical Clinic 1 for Internal Medicine under the supervision of a professional dialysis unit.

### Preparation of PBMCs

Peripheral blood was collected in S-Monovette K3 EDTA blood collection tubes (Sarstedt, Germany). Collected blood was pre-diluted in PBS/BSA (Gibco, USA) at a 1:1 ratio and underlaid with 15 mL Ficoll-Paque Plus (GE Healthcare, USA). Tubes were centrifuged at 800 g for 20 minutes at room temperature. Isolated PBMC were washed twice with PBS/BSA and stored at −80°C until use as previously described^65^.

### Antigen stimulation of T cells

Isolated PBMC were stimulated with 1µg/ml Sars-CoV-2 OPPs (Miltenyi, Germany), 1 µg/mL β1+β2 or 1 µg/ml M3+M4 purified cell lysates (Biotrend, Germany). 2.5×10^6^ PBMCs were plated for each condition in 96-UWell Plates in RPMI media (Life Technologies), supplemented with 1% Penicillin-Streptomycin-Glutamine (Sigma Aldrich, USA), and 10% FCS (PAN-Biotech, USA) and were stimulated or left untreated as a control for 16 hours. As a positive control, cells were stimulated with SEB (1 µg/ml, Sigma Aldrich, USA) or left untreated as a negative control. After 2 hours, Brefeldin A (1µg/ml, Sigma Aldrich, USA) was added. As previously applied by our groups and others, antigen-specific responses were considered positive after the non-specific background was subtracted, and more than 0.001% or at least 15 positive cells were detectable. Negative values were set to zero.

### Measurement of autoantibodies to GPCR

Serum was collected in a S-Monovette Serum collection tube (Sarstedt, Germany) and processed per manufacturer’s instructions. Purified serum was stored at −20 °C until use. ELISA systems for detection of autoantibodies against β-1 Adrenergic Receptor (β1), β-2 Adrenergic Receptor (β2), Muscarinergic Choline Receptor 3 (M3), and Muscarinergic Choline Receptor 4 (M4) were all obtained from CellTrend, Germany. Autoantibodies in serum were determined per manufacturer’s instructions and normalized using the provided standard. The following cut-offs were used to define presence of autoantibodies: β1: >15 U/ml; β2: >8 U/ml; M3: >6 U/ml; M4: >10.7 U/ml.

### QPCR measurement of EBV virus titer

Peripheral blood samples were monitored for EBV by qPCR. DNA was isolated from whole blood samples using AltoStar Purification Kit 1.5 (Altostar, Germany) and qPCR was performed using AltoStar EBV PCR Kit 1.5 following the manufacturer’s instructions. Both Kits were performed on an AltoStar AM16 pipetting robot and qPCR was measured on an BioRad C1000 Thermal Cycler. The detection level was determined as the lowest viral load measured within the range of linearity (250 copies/ml).

### Cytokine analysis

Purified serum was stored at −20 °C until use. The level of the cytokines IL-4, IL-2, IP-10 (CXCL10), IL-1β, TNF-α, MCP-1 (CCL2), IL-17A, IL-6, IL-10, IFN-γ, IL-12p70, IL-8 (CXCL8), and TGF-β1 (free active) was assessed using the LEGENDplex Human Essential Immune Response Panel (BioLegend, San Diego, USA) per manufacturers instruction and acquired on a CytoFLEX (Beckman Coulter, Germany) flow cytometer. Briefly, serum was incubated with beads coated with antibodies for one of the analyzed cytokines together with a secondary antibody and phycoerythrin-conjugated (PE) detection antibody. Each cytokine is detectable by the bead size and level of allophycocyanin (APC) within the bead, and the intensity of PE is relative to the levels of the cytokine in the sample. The measured fluorescence intensities were normalized using the provided cytokine standard.

### ME/CFS symptoms and neuropsychological evaluation

Long-COVID symptoms were quantified using a 19 items questionnaire based on the ISARIC COVID-19 follow-up study protocol. To quantify the severity of mental and physical fatigue, all patients completed the Chalder fatigue scale, Munich Berlin Symptom Questionnaire (MBSQ) and Epworth Sleepiness Scale (ESS). Physical function and impairment were assessed by the Bell disability scale and the Short Form Health Survey-36 (SF-36). Additionally, hand grip strength was measured as an objective parameter for physical impairment using a hydraulic hand dynamometer (Saehan, Germany). Furthermore, dysautonomic symptoms were evaluated using the COMPASS-31 score. All neuropsychological assessments were conducted using the German version of the “Consortium to Establish a Registry for Alzheimer’s Disease Neuropsychological Assessment Battery” (CERAD). The standard assessment, which includes evaluation of verbal fluency (animal naming), a modified version of the Boston Naming Test, global cognition (Mini-Mental State examination), verbal memory (word list learning, delayed recall), as well as constructional practice and delayed recall, was widen out by additional items on processing speed (Trail Making Test A) and executive functioning (Trail Making Test B, letter fluency: S-words).

### Spike Protein Elisa

Serum was collected in a S-Monovette Serum collection tube (Sarstedt, Germany) and stored at −20 °C until use. The concentration of spike protein was assessed using the Human SARS-CoV.2 RBD Elisa Kit (Invitrogen, Thermo Fisher, USA). The Elisa was performed according to the manufacturers technical manual and no alterations were implemented. The optical density of the plates was determined using a Asys UVM 340.

### Antibodies

Antigen specific T cells (all antibodies are from BioLegend (USA) unless otherwise noted): Surface staining: CCR7 (CD197)-PerCP-Cy5.5; clone: G043H7, CD4-A700; clone: OKT4, LD eFluor780 (eBioscience, USA), CD8-V500; clone: RPA-T8 (BD Biosciences), CD45RA-BV605; clone: HI100. Intracellular staining: CD137 (4-1BB)-PE-Cy7; clone: 4B4-1, CD154 (CD40L)-A647; clone: 24-31, TNFa-eFluor450; clone: MAb11 (eBioscience, USA), CD3-BV785; clone: OKT3, PD-1-PE; clone: A17188A, Tim-3-VioBright-Fitc; clone: REA635 (Miltenyi, Germany), CTLA-4-BV421; clone: BNI3, Lag-3-BV650; clone: 11C3C65. Fixable Viability Dye eFluor 780 (eBioscience, USA) was used for live/dead discrimination.

### Flow Cytometry

Stimulated PBMC were extracellular stained with optimal concentrations of antibodies for 10 minutes at room temperature in the dark. Cells were washed twice with PBS/BSA before preparation for intracellular staining using the Intracellular Fixation & Permeabilization Buffer Set (Thermo Fisher Scientific) as per manufacturer’s instructions. Fixed and permeabilized cells were stained for 30 minutes at room temperature in the dark with an optimal dilution of antibodies against the intracellular antigen.

All samples were immediately acquired on a CytoFlex flow cytometer (Beckman Coulter). Quality control was performed daily using the recommended CytoFlex Daily QC Fluorospheres (Beckman Coulter). No modification to the compensation matrices was required throughout the study. Flow cytometry data were analyzed using FlowJo version 10.6.2 (BD Biosciences); gating strategies are presented in Supplemental Figures S5+S6.

### Statistical analysis

Statistical analysis was performed using R, version 4.2.1. Categorical variables are summarized as numbers and frequencies; quantitative variables are reported as mean ± SD. No data transformation was used. Violin plots depict the median and the first and third quartiles. The whiskers correspond to 1.5 times the interquartile range. All applied statistical tests are paired and two-sided. Dotted lines between violins indicate the individual patients. Differences in quantitative variables between all three groups are analyzed using non-parametric Wilcoxon test. P values below 0.050 were considered significant. P values were not corrected for multiple testing, as this study was of exploratory nature^66,67^. The normalization standard for the ELISA results was calculated using a four parameter logistic regression. Parameter correlation was assessed using Spearman’s rank correlation coefficient.

## Data Availability

All data produced in the present study are available upon reasonable request to the authors

## Author contributions

Conceptualization: T.W. and N.B.; Data curation: M.A., L.W., K.R. and U.S.; Formal analysis: M.A., L.W., K.R. and U.S.; Funding acquisition: N.B.; Investigation: M.A., L.W., K.R., S.S., J.K., S.K., U.S. and N.B.; Methodology: M.A., U.S. and N.B.; Resources: A.D., F.S. and T.W.; Visualization: M.A. and K.R.; Writing – original draft: M.A., L.W., K.R., U.S. and N.B.; Writing - review & editing: M.A., L.W., K.R., U.S. and N.B.;

## Acknowledgments

We want to express our deepest gratitude to the patients who donated their blood samples and clinical data for this project. We thank Miltenyi Biotec who supported our study by providing materials and equipment (TheraSorb columns) for Immunoadsorption. This work was further supported by DFG grants NFDI4 (501875662)

## Conflict of interest

The study was initiated by the investigators themselves. They independently created the study without any external guidance or involvement from entities like Miltenyi Biotec or any other undisclosed parties, pertaining to its design, analysis, data interpretation or manuscript approval. The authors themself did not receive any payment related to this study and have stated that they have no conflicting interests.

## Notes

### Competing Interest Statement

The authors have declared no competing interest.

### Author Declarations

The study was approved by the ethical committee of Ruhr university Bochum (Reg.-Nr. 23-7752) and all participants provided a written informed consents.

