## Supplemental Materials for "Effect of Immunoadsorption on clinical presentation and immune alterations in COVID-19–associated ME/CFS"

**Table S1**

Percentage of Autoantibody-Reconstution 30 days after IA. Percent is calculated to levels before IA

| PID | β1 | β2 | M3 | M4 |
|---|---|---|---|---|
| IA01 | 107,72% | 94,79% | 101,99% | 98,93% |
| IA02 | 49,17% | 76,44% | 92,81% | 85,80% |
| IA03 | 107,61% | 64,92% | 86,22% | 144,00% |
| IA04 | 79,34% | 73,76% | 80,55% | 40,69% |
| IA05 | 91,47% | 145,02% | 177,06% | 177,17% |
| IA06 | 111,51% | 64,78% | 67,57% | 64,18% |
| IA07 | 180,29% | 25,79% | 38,20% | 31,00% |
| IA08 | 55,28% | 180,48% | 176,75% | 214,69% |
| IA10 | 96,37% | 61,54% | 75,30% | 69,53% |
| IA11 | 54,10% | 144,72% | 49,41% | 45,61% |
| IA12 | 98,14% | 75,85% | 84,18% | 91,69% |
| IA13 | 68,26% | 62,39% | 76,31% | 139,29% |
| Median | 93,92% | 74,81% | 82,36% | 88,75% |

β1 = β1-adrenergic receptor; β2 = β2-adrenergic receptor; M3 = M3-mACh receptor; M4 = M4-mACh receptor.

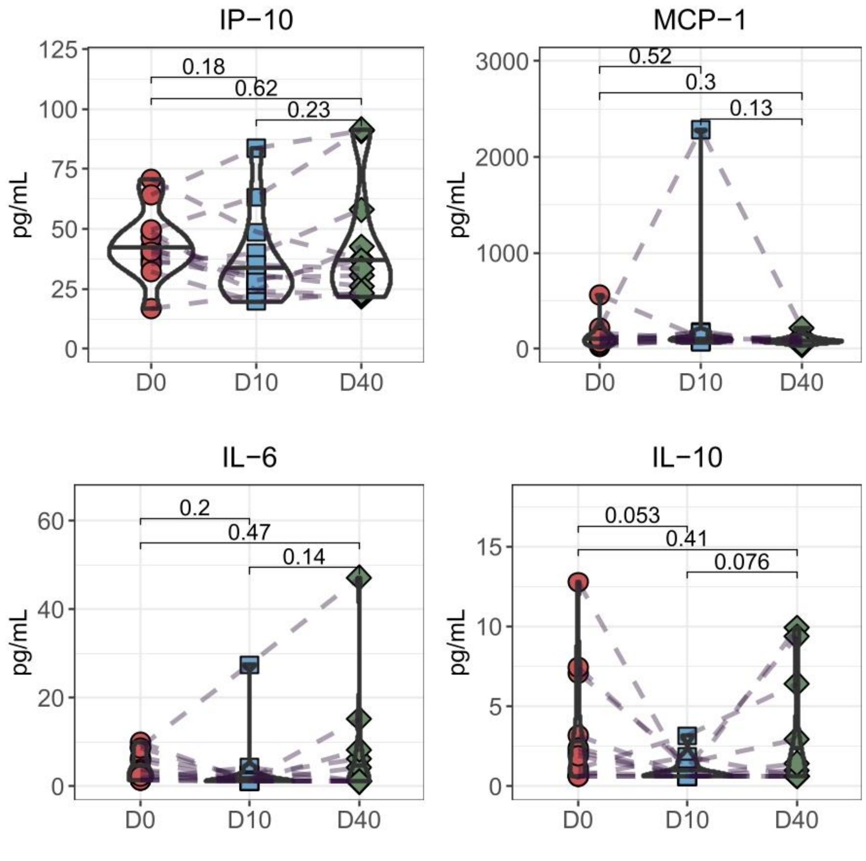

**Supplemental Figure S1.** Legendplex analysis of the IP-10, MCP-1 , IL-6 and IL-10 cytokine profile of ME/CSF patients before IA (D0), immediately after IA (D10) and 30 days after the last IA (D40). Pairwise Man-Whitney U Test was performed.

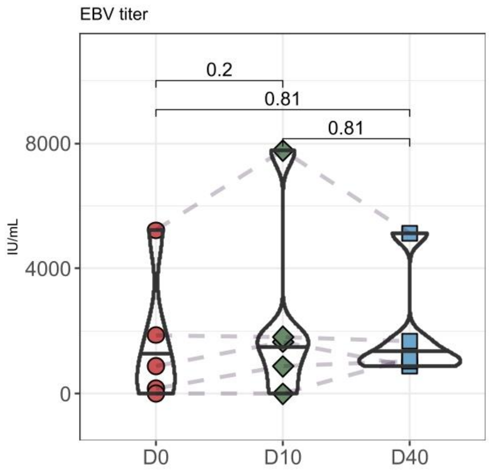

**Supplemental Figure S2.** QPCR analysis of EBV virus titer in ME/CSF patients before IA (D0), immediately after IA (D10) and 30 days after the last IA (D40). Pairwise Man-Whitney U Test was performed.

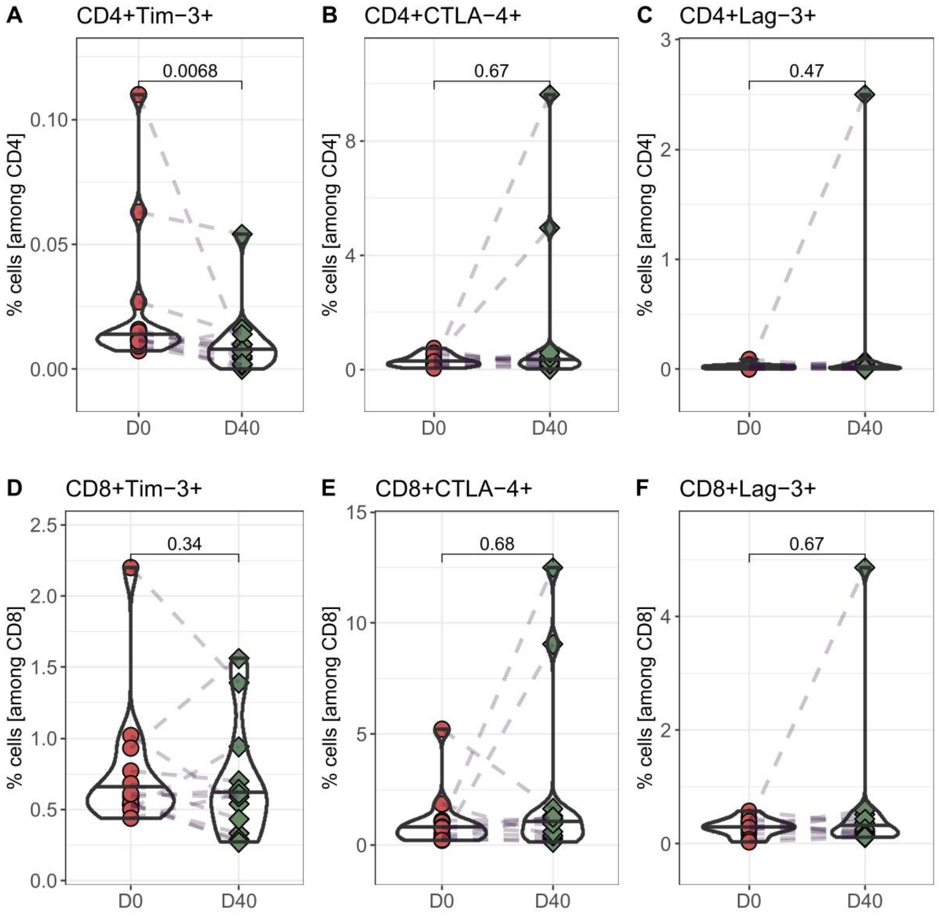

**Supplemental Figure S3.** Analysis of exhaustion marker on T cells in ME/CSF patients. PBMCs from patients were isolated before IA (D0) and 30 days after the last IA (D40) and stained with the appropriate antibodies and analyzed in a flow-cytometer. T cells were identified as $CD3^+$ and $CD4^+$ or $CD8^+$ lymphocytes and cells were stained with the respective exhaustion markers. Pairwise Man-Whitney U Test was performed.

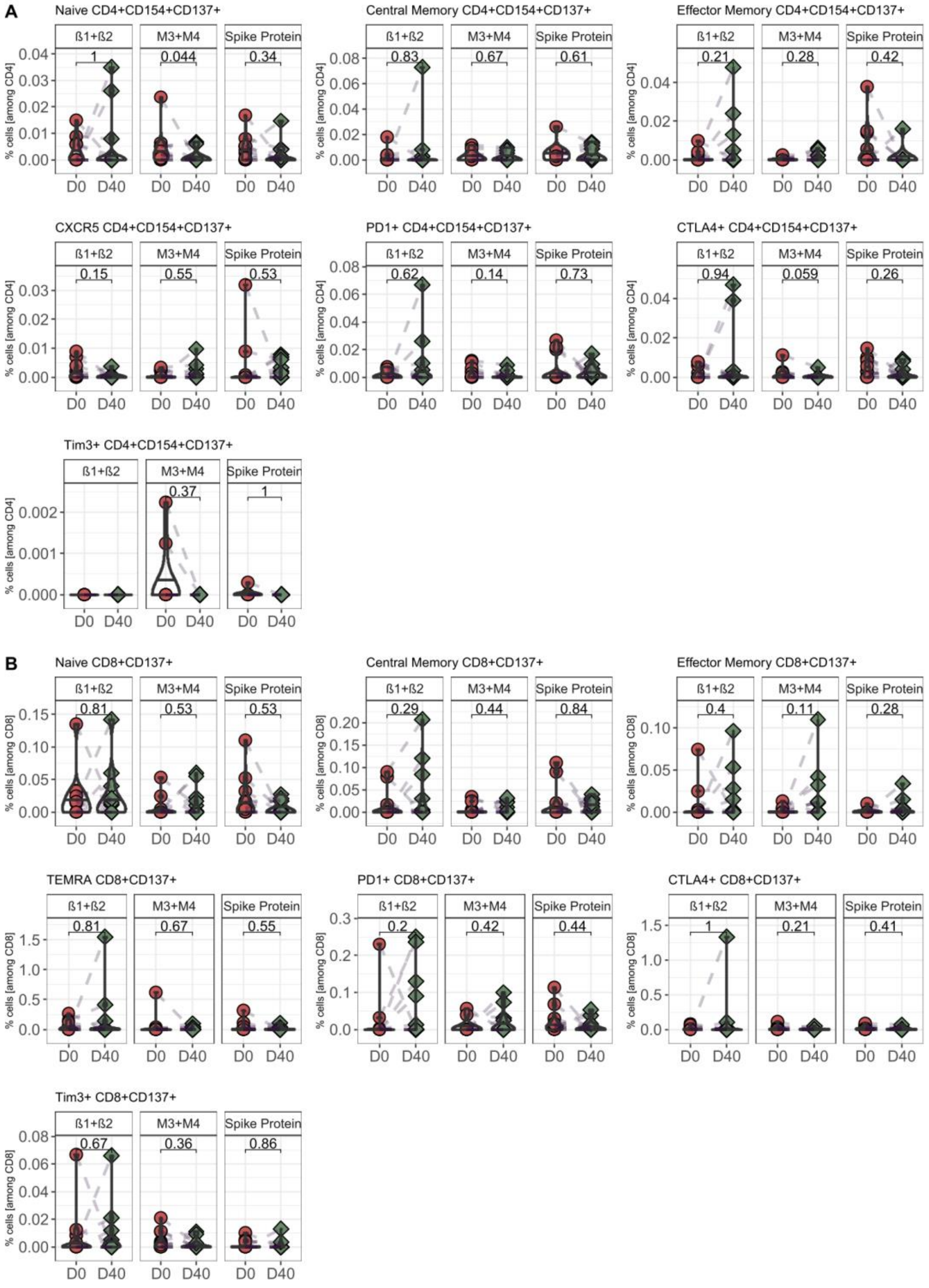

**Supplimental Figure S4. Frequenz of SARS-CoV-2 spike protein specific CD4 (A) and CD8 (B) T cells.** PBMCs from patients were isolated before IA (D0) and 30 days after the last IA (D40) and stained with the appropriate antibodies and analyzed in a flow-cytometer. T cells were identified as $CD3^+$ and $CD4^+$ or $CD8^+$. Naïve and memory T cells were distingished by expression of CD45RA and CCR7 and Exhausted T cells by expression of PD-1, Tim-3 and CTLA-4. PBMC were stimulate for 18h with SARS-CoV-2 Spike Protein overlapping peptideso or β1+β2 / M3+M4 GCPR cell lysates, stained with the appropriate antibodies and analyzed in a flow-cytometer. Activated T helper cells were identified as $CD4^+$ $CD154^+CD137^+$ and activated cytotoxic T cells as $CD8^+CD137^+$. Pairwise Man-Whitney U Test was performed. Gating strategies are depicted in supplementary figure S5+S6.

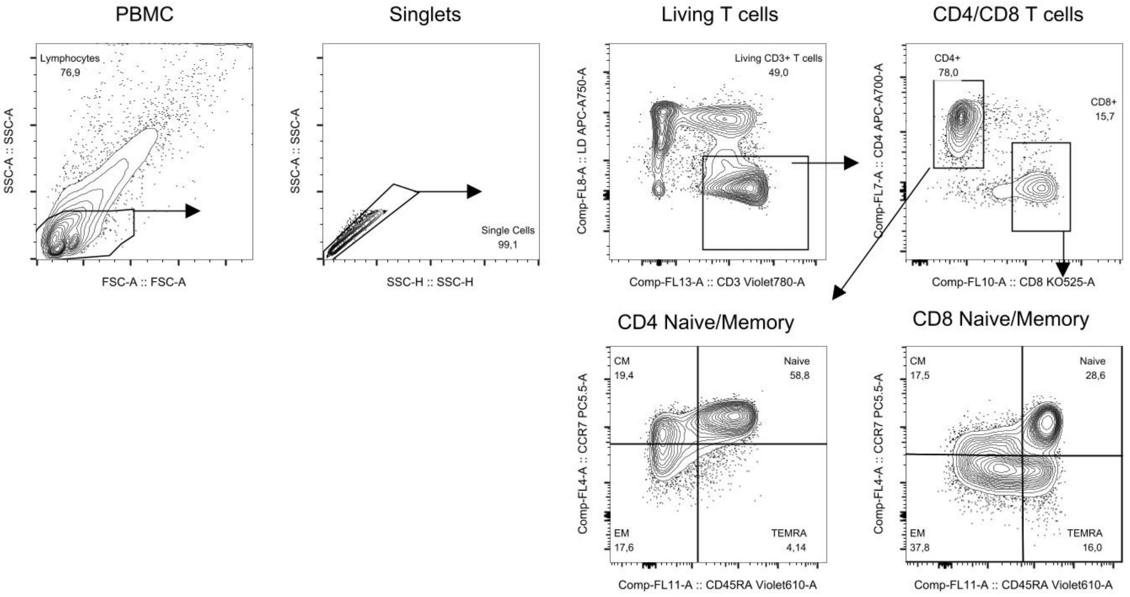

**Supplemental Figure S5.** Gating strategy to identify T cell subpoulations. Lymphocytes were identified by SSC/FSC-profile. Doublets and dead cells were excluded and T cells were identified as $CD3^+$ and $CD4^+$ or $CD8^+$ lymphocytes. Naïve T cells were identified as $CD45RA^+CCR7^+$ and memory T cells were combined of central memory T cells (CM, $CD45RA^-CCR7^+$), effector memory T cells (EM, $CD45RA^-CCR7^-$) and TEMRA cells ($CD45RA^+CCR7^-$).

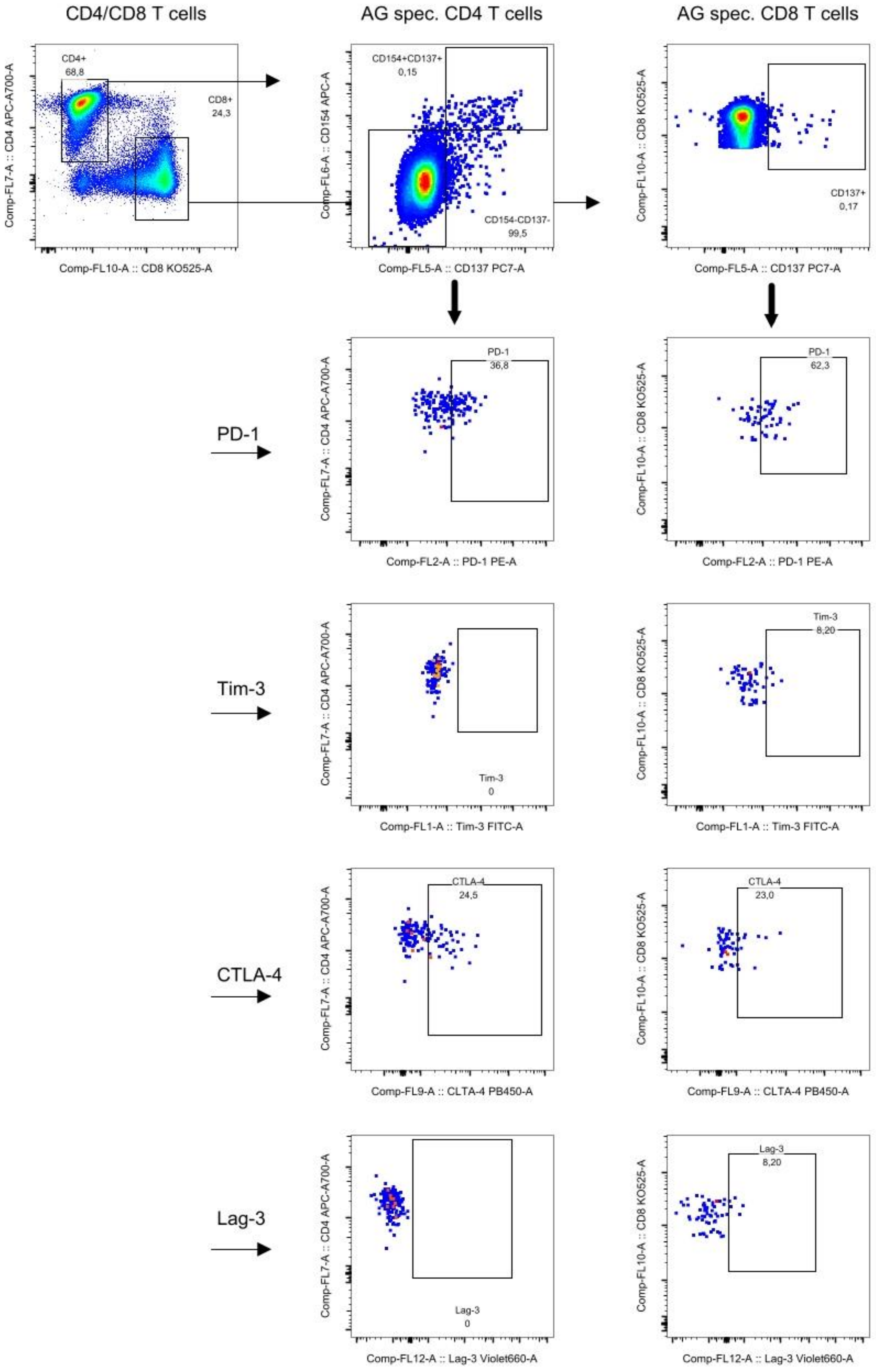

**Supplemental Figure S6.** Gating strategy to identify SARS-CoV-2 spike protein or GCPR-specific T cell subpoulations. PBMC were stimulated for 18h with β1/β2-adrenoreceptors or muscarinic M3/M4 receptor cell lysates or SARS-CoV-2 spike protein OPPs and activated T helper cells were identified as $CD4^{+}$ $CD154^{+}CD137^{+}$ and activated cytotoxic T cells as $CD8^{+}CD137^{+}$. T cells were stained with antibodies against exhaustion marker PD-1, Tim-3, CTLA-4 and Lag-3 and analyzed in a flow-cytometer.
